# Excess mortality associated with the COVID-19 pandemic among Californians 18–65 years of age, by occupational sector and occupation: March through October 2020

**DOI:** 10.1101/2021.01.21.21250266

**Authors:** Yea-Hung Chen, Maria Glymour, Alicia Riley, John Balmes, Kate Duchowny, Robert Harrison, Ellicott Matthay, Kirsten Bibbins-Domingo

## Abstract

**Background:** Though SARS-CoV-2 outbreaks have been documented in occupational settings and though there is speculation that essential workers face heightened risks for COVID-19, occupational differences in excess mortality have, to date, not been examined. Such information could point to opportunities for intervention, such as workplace modifications and prioritization of vaccine distribution.

**Methods and findings:** Using death records from the California Department of Public Health, we estimated excess mortality among Californians 18–65 years of age by occupational sector and occupation, with additional stratification of the sector analysis by race/ethnicity. During the COVID-19 pandemic, working age adults experienced a 22% increase in mortality compared to historical periods. Relative excess mortality was highest in food/agriculture workers (39% increase), transportation/logistics workers (28% increase), facilities (27%) and manufacturing workers (23% increase). Latino Californians experienced a 36% increase in mortality, with a 59% increase among Latino food/agriculture workers. Black Californians experienced a 28% increase in mortality, with a 36% increase for Black retail workers. Asian Californians experienced an 18% increase, with a 40% increase among Asian healthcare workers. Excess mortality among White working-age Californians increased by 6%, with a 16% increase among White food/agriculture workers.

**Conclusions:** Certain occupational sectors have been associated with high excess mortality during the pandemic, particularly among racial and ethnic groups also disproportionately affected by COVID-19. In-person essential work is a likely venue of transmission of coronavirus infection and must be addressed through strict enforcement of health orders in workplace settings and protection of in-person workers. Vaccine distribution prioritizing in-person essential workers will be important for reducing excess COVID mortality.

## Introduction

More deaths are occurring during the COVID-19 pandemic than predicted by historical trends [1-4]. In California, per-capita excess mortality is relatively high among Blacks, Latinos, and individuals with low educational attainment [4]. An explanation for these findings is that these populations face unique occupational risks because they may disproportionately make up the state’s essential workforce and because essential workers often cannot work from home [4-6]. Additionally, due to historical structural inequities, low-wage essential workers may be more likely to live in crowded housing [5-7], resulting in household transmission.

Despite the inherent risks that essential workers face, no study to date has examined differences in excess mortality across occupation. Such information could point to opportunities for intervention, such as workplace modifications and prioritization of vaccine distribution. Using time-series models to forecast deaths from March through October 2020, we compare excess deaths among California residents 18–65 years of age across occupational sectors and occupations, with additional stratification of the sector analysis by race/ethnicity.

## Methods

We obtained data from the California Department of Public Health on all deaths occurring on or after January 1, 2016.

To focus on individuals whose deaths were most plausibly linked to work, we restricted our analysis to decedents 18–65 years of age. Death certificates include an open text field for “Decedent’s usual occupation,” described as “type of work done during most of working life.” Retirement is not separately recorded. We processed the occupation information listed on the death certificates using an automated system developed by the National Institute for Occupational Safety and Health, which converts free-text occupational data to 2010 US Census codes. A team of 3 researchers manually categorized the resulting 529 unique codes into occupational sectors, with a focus on the 13 sectors identified by Cailfornia officials as comprising the state’s essential workforce[8] and retail workers; we anticipated that these sectors would be most at risk. To ease presentation, we combined or eliminated some sectors, placing the defense, communications/IT, and financial sectors in the not-essential category (under the logic that it was particularly difficult to ascertain which workers in these sectors fully met the state’s definitions for essential work) and placing chemical, energy, and water sectors in the facilities category. This resulted in the following 9 groups: facilities, food/agriculture, government/community, health/emergency, manufacturing, retail, transportation/logistics, not essential, and unemployed/missing. We defined 4 racial/ethnic groups: Asian, Black, Latino, and White, with the definition of Latino overwriting any racial designation in the death records. Our definition of Asian, Black, and White excludes individuals identified on the death certificate as multiracial.

We defined pandemic time as beginning on March 1, 2020. In some time-stratified analysis, we compared the months of March through May to the months of June and July. We chose the cutoff of June 1 because it is roughly 3 weeks after the state’s post-shutdown reopening in early May, and because we anticipate lags between policy, infection, and death. Similarly, the ending date of July 31 is roughly 3 weeks after the state ordered restaurants and indoor businesses to close in early July.

We conducted time-series analysis for each occupational sector, with additional stratification by race/ethnicity. For each group of interest (for example, each occupational sector of interest), we repeated the following procedure. We aggregated the data to months or weeks, using the weekly analysis for visualizations and the monthly analysis to derive summary measures. Following our previous work [4], we fit dynamic harmonic regression models with autoregressive integrated moving average (ARIMA) errors for the number of monthly/weekly all-cause deaths, using deaths occurring among the group prior to March 1, 2020. For each iteration, we used a model-fitting procedure described by Hyndman and Khandakar [9]. Using the final model, we forecast the number of deaths for each unit of time, along with corresponding 95% prediction intervals (PI). To obtain the total number of excess deaths for the entire time window, we subtracted the total number of expected (forecast) deaths from the total number of observed deaths. We obtained a 95% PI for the total by simulating the model 10,000 times, selecting the 97.5% and 2.5% quantiles, and subtracting the total number of observed deaths. We report in our tables the observed number of deaths divided by the expected number of deaths, as predicted by our models. We interpret these ratios as risk ratios for mortality, comparing pandemic time to non-pandemic time. We also estimated excess mortality for all specific occupations; for individual occupations, we defined excess mortality and risk ratios by comparing 2020 deaths to the arithmetic mean of 2018 and 2019 deaths.

We conducted all analyses in R, version 4.04.

## Results

We estimate that from March 2020 through October 2020, there were 10,047 (95% PI: 9,229–10,879) excess deaths among Californians 18–65 years of age (Table 1). Relatively large numbers of excess deaths were recorded among workers in the facilities sector (1,681; 95% PI: 1,447–1,919) and the transportation/logistics sector (1,542; 95% PI: 1,350–1,738). Relative to pre-pandemic time, mortality increased during the pandemic by 39% among food/agriculture workers (risk ratio RR=1.39; 95% PI: 1.32–1.48), 28% among transportation/logistics workers (RR=1.28; 95% PI: 1.24–1.33), 27% among facilities workers (RR=1.27; 95% PI: 1.22–1.32), and 23% (RR=1.23; 95% PI: 1.18–1.28) among manufacturing workers.

**Table 1.** Excess mortality among Californians 18–65 years of age, by occupational sector: March through October 2020.

|  | Excess deaths | Risk ratio<sup>a</sup> |
| --- | --- | --- |
| Entire state | 10,047 (9,229–10,879) | 1.22 (1.20–1.24) |
| Facilities | 1,681 (1,447–1,919) | 1.27 (1.22–1.32) |
| Food or agriculture | 1,050 (897–1,204) | 1.39 (1.32–1.48) |
| Government or community | 422 (324–520) | 1.14 (1.11–1.18) |
| Health or emergency | 585 (523–647) | 1.19 (1.17–1.22) |
| Manufacturing | 638 (530–749) | 1.23 (1.18–1.28) |
| Retail | 646 (517–778) | 1.18 (1.14–1.23) |
| Transportation or logistics | 1,542 (1,350–1,738) | 1.28 (1.24–1.33) |
| Not essential | 1,167 (910–1,428) | 1.11 (1.08–1.14) |
| Unemployed or missing | 1,969 (1,718–2,225) | 1.23 (1.19–1.27) |

<sup>a</sup> Risk ratios are defined as the observed number of deaths divided by the expected number of deaths. They are interpretable as the risk ratio for mortality, comparing pandemic time to non-pandemic time.

Relative increases in mortality varied over time (Fig 1) and by occupational sector (Fig 2). In March through May, there was a 14% increase in mortality among all working-age Californians (RR=1.14; 95% PI: 1.09–1.20) compared to a 31% increase among workers in the food/agriculture (RR=1.31; 95% PI: 1.17–1.49). In the months of June and July, the RR were particularly high in the food/agriculture (RR=1.61; 95% PI: 1.44–1.83), transportation/logistics (RR=1.52; 95% PI: 1.38–1.69), manufacturing (RR=1.52; 95% PI: 1.37–1.72), and facilities sectors (RR=1.44; 95% PI: 1.31–1.61).

**Figure 1.**
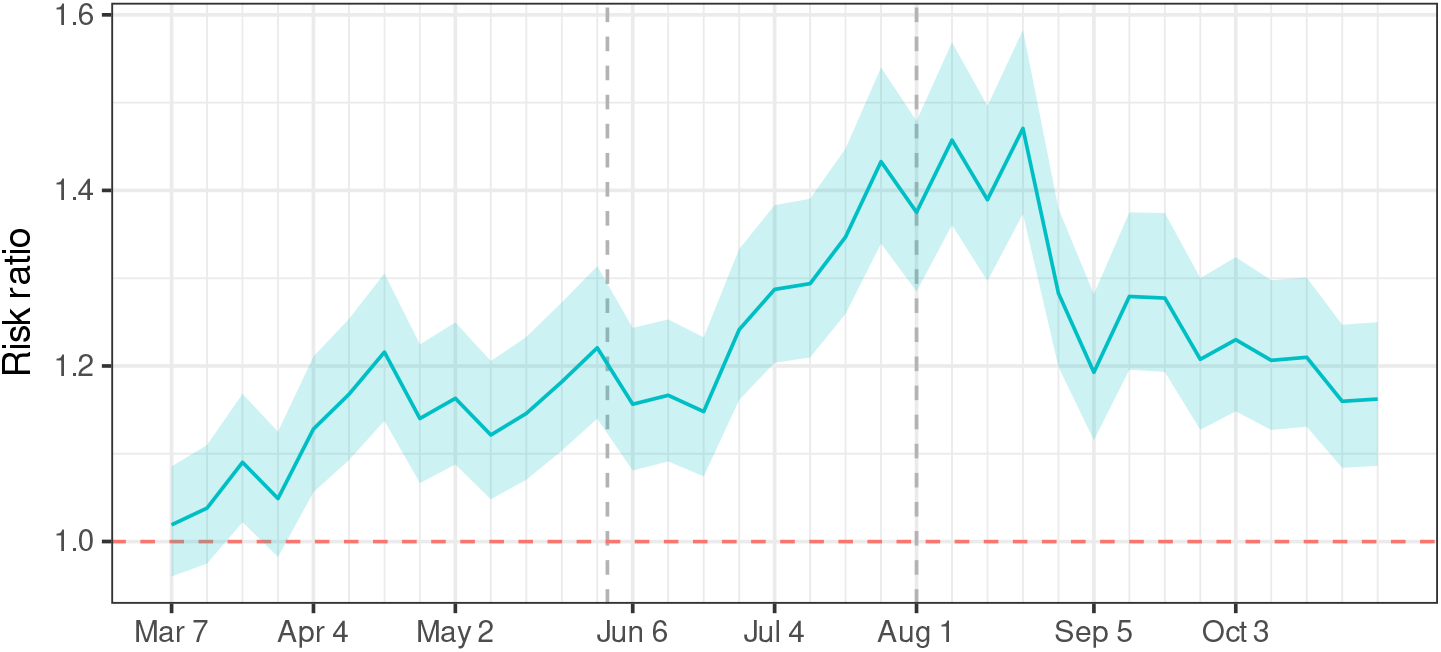
Risk ratios for death, comparing pandemic time to non-pandemic time, among Californians 18–65 years of age, March through October 2020. The dashed vertical lines mark boundaries between phases of California’s major pandemic policies, lagged to acknowledge time from policy decisions to infection to death. The first phase corresponds to a period of sheltering in place, while the second phase corresponds to a period of reopening.

**Figure 2.**
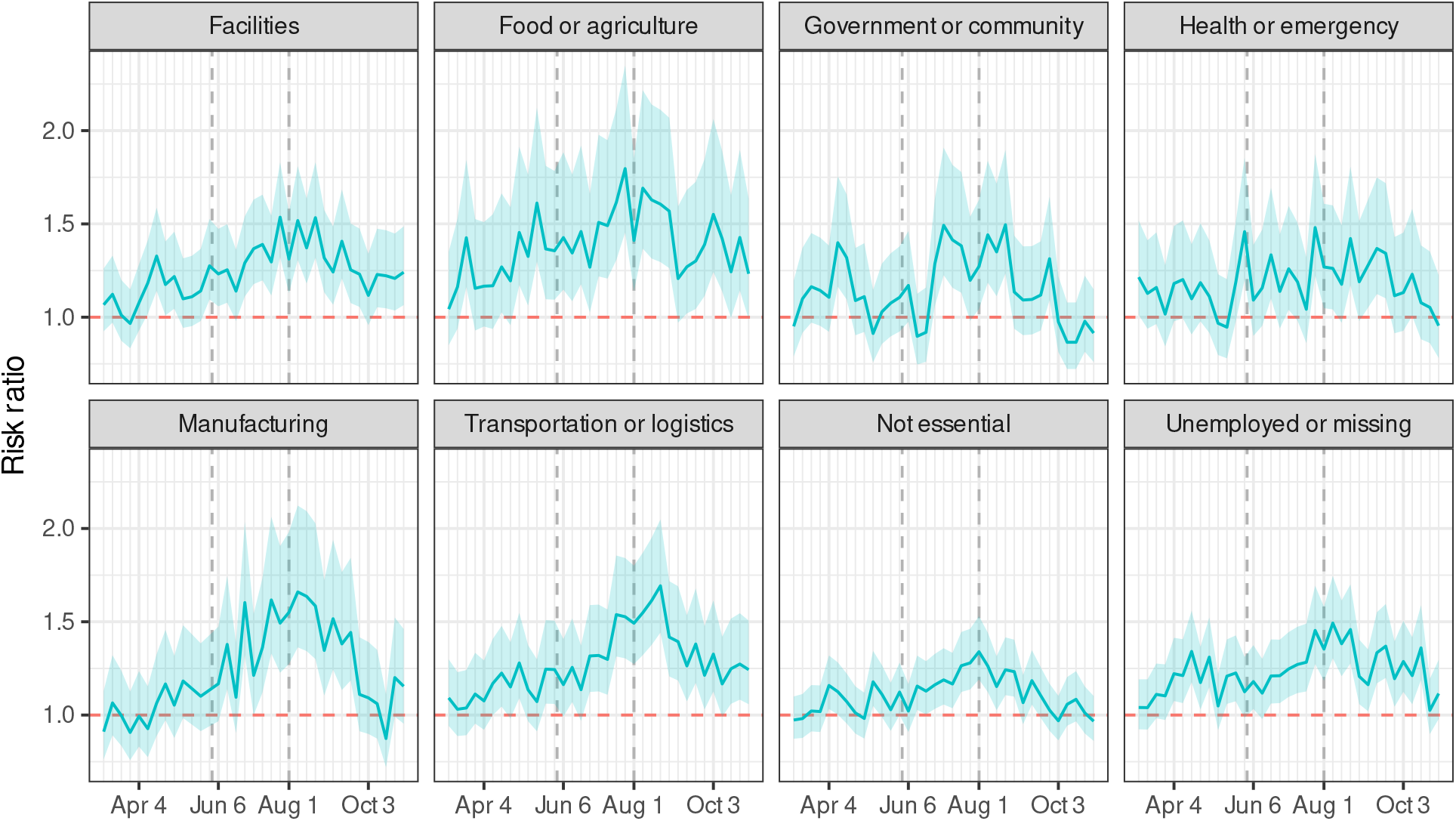
Risk ratios for death, comparing pandemic time to non-pandemic time, among Californians 18–65 years of age, by occupational sector, March through October 2020. The dashed vertical lines mark boundaries between phases of California’s major pandemic policies, lagged to acknowledge time from policy decisions to infection to death. The first phase corresponds to a period of sheltering in place, while the second phase corresponds to a period of reopening.

RR also varied by race/ethnicity (Table 2). Latino Californians experienced a 36% increase in mortality during the pandemic (RR=1.36; 95% PI: 1.29–1.44), with a 59% increase among Latino food/agriculture workers (RR=1.59; 95% PI: 1.47–1.75). Black Californians experienced a 28% increase in mortality (RR=1.28; 95% PI: 1.24–1.33), with a 36% increase for Black retail workers (RR=1.36; 95% PI: 1.21–1.55). Asian Californians experienced an 18% increase (RR=1.18; 95% PI: 1.14–1.23), with a 40% increase among Asian healthcare workers (RR=1.40; 95% PI: 1.33–1.49). Mortality among White working-age Californians increased by 6% (RR=1.06; 95% PI: 1.02–1.12) with a 16% increase among White food/agriculture workers (RR=1.16; 95% PI: 1.09–1.24).

**Table 2.** Risk ratios for mortality, comparing pandemic time to non-pandemic time, among California residents 18–65 years of age, by occupational sector and race/ethnicity, March through October 2020.

|  | All races | Asian | Black | Latino | White |
| --- | --- | --- | --- | --- | --- |
| All sectors | 1.22 (1.20–1.24) | 1.18 (1.14–1.23) | 1.28 (1.24–1.33) | 1.36 (1.29–1.44) | 1.06 (1.02–1.12) |
| Food or agriculture | 1.39 (1.32–1.48) | 1.18 (1.05–1.33) | 1.34 (1.19–1.54) | 1.59 (1.47–1.75) | 1.16 (1.09–1.24) |
| Transportation or logistics | 1.28 (1.24–1.33) | 1.26 (1.12–1.44) | 1.35 (1.26–1.46) | 1.40 (1.31–1.52) | 1.10 (1.02–1.20) |
| Facilities | 1.27 (1.22–1.32) | 1.24 (1.08–1.46) | 1.25 (1.17–1.34) | 1.38 (1.27–1.51) | 1.11 (1.04–1.20) |
| Unemployed or missing | 1.23 (1.19–1.27) | 1.08 (1.04–1.14) | 1.31 (1.22–1.40) | 1.31 (1.22–1.41) | 1.09 (1.01–1.20) |
| Manufacturing | 1.23 (1.18–1.28) | 1.18 (1.06–1.33) | 1.13 (1.01–1.30) | 1.44 (1.34–1.57) | 1.00 (0.92–1.10) |
| Health or emergency | 1.19 (1.17–1.22) | 1.40 (1.33–1.49) | 1.27 (1.17–1.40) | 1.32 (1.18–1.51) | 1.02 (0.96–1.10) |
| Retail | 1.18 (1.14–1.23) | 1.10 (1.00–1.22) | 1.36 (1.21–1.55) | 1.40 (1.28–1.55) | 1.08 (1.04–1.13) |
| Government or community | 1.14 (1.11–1.18) | 1.22 (1.07–1.41) | 1.20 (1.09–1.33) | 1.42 (1.32–1.53) | 0.96 (0.89–1.04) |
| Not essential | 1.11 (1.08–1.14) | 1.14 (1.06–1.23) | 1.23 (1.15–1.33) | 1.29 (1.20–1.41) | 1.00 (0.95–1.07) |

Per occupation (Table 3), risk ratios for mortality comparing pandemic time to non-pandemic time were highest among cooks (RR=1.60), packaging and filling machine operators and tenders (RR=1.59), miscellaneous agricultural workers (RR=1.55), bakers (RR=1.50), and construction laborers (RR=1.49).

**Table 3.**
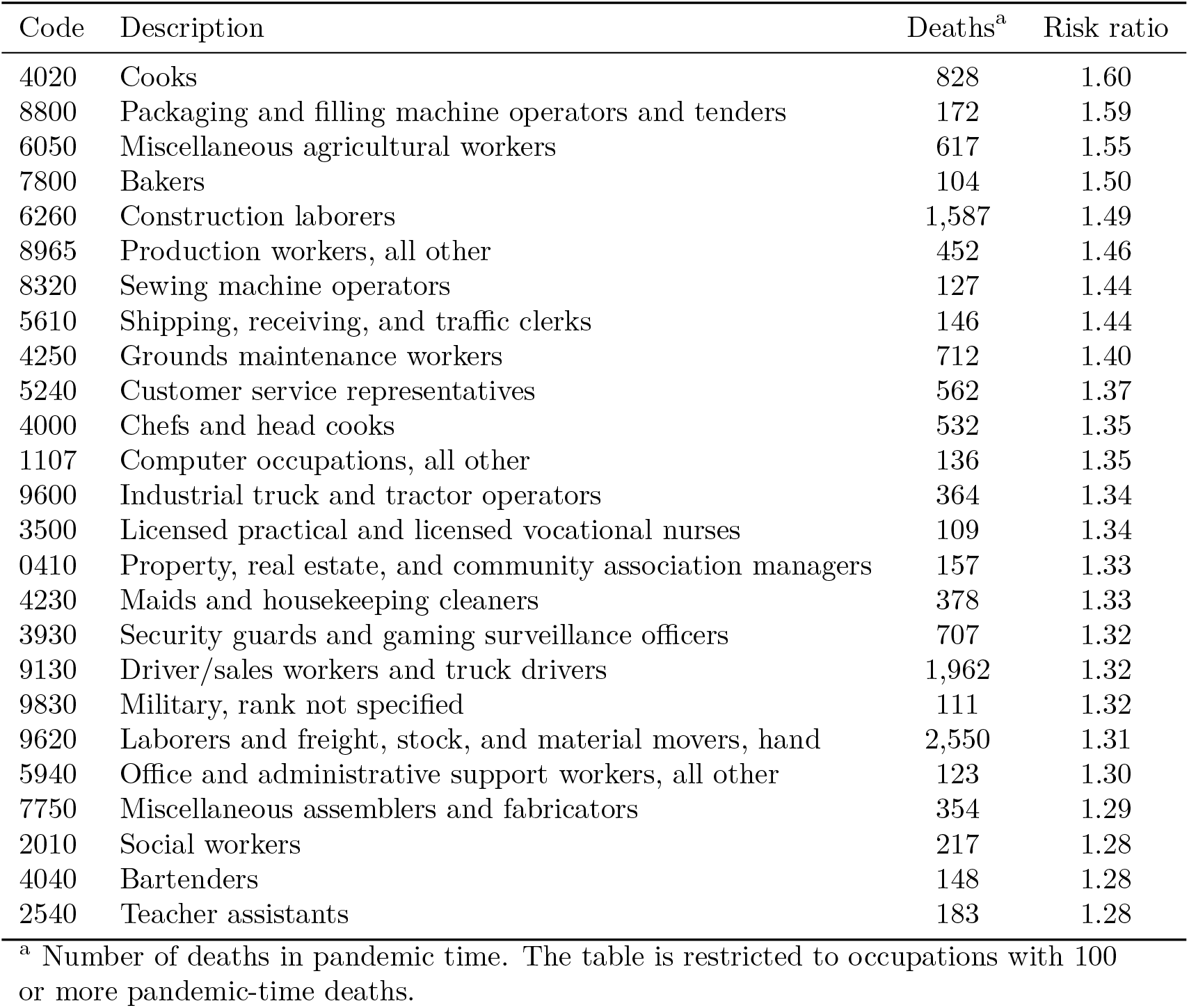
Risk ratios for mortality, comparing pandemic time to non-pandemic time, among California residents 18–65 years of age, by occupation, March through October 2020.

| Code | Description | Deaths<sup>a</sup> | Risk ratio |
| --- | --- | --- | --- |
| 4020 | Cooks | 828 | 1.60 |
| 8800 | Packaging and filling machine operators and tenders | 172 | 1.59 |
| 6050 | Miscellaneous agricultural workers | 617 | 1.55 |
| 7800 | Bakers | 104 | 1.50 |
| 6260 | Construction laborers | 1,587 | 1.49 |
| 8965 | Production workers, all other | 452 | 1.46 |
| 8320 | Sewing machine operators | 127 | 1.44 |
| 5610 | Shipping, receiving, and traffic clerks | 146 | 1.44 |
| 4250 | Grounds maintenance workers | 712 | 1.40 |
| 5240 | Customer service representatives | 562 | 1.37 |
| 4000 | Chefs and head cooks | 532 | 1.35 |
| 1107 | Computer occupations, all other | 136 | 1.35 |
| 9600 | Industrial truck and tractor operators | 364 | 1.34 |
| 3500 | Licensed practical and licensed vocational nurses | 109 | 1.34 |
| 0410 | Property, real estate, and community association managers | 157 | 1.33 |
| 4230 | Maids and housekeeping cleaners | 378 | 1.33 |
| 3930 | Security guards and gaming surveillance officers | 707 | 1.32 |
| 9130 | Driver/sales workers and truck drivers | 1,962 | 1.32 |
| 9830 | Military, rank not specified | 111 | 1.32 |
| 9620 | Laborers and freight, stock, and material movers, hand | 2,550 | 1.31 |
| 5940 | Office and administrative support workers, all other | 123 | 1.30 |
| 7750 | Miscellaneous assemblers and fabricators | 354 | 1.29 |
| 2010 | Social workers | 217 | 1.28 |
| 4040 | Bartenders | 148 | 1.28 |
| 2540 | Teacher assistants | 183 | 1.28 |

<sup>a</sup> Number of deaths in pandemic time. The table is restricted to occupations with 100 or more pandemic-time deaths.

## Discussion

Our analysis of deaths among Californians between the ages of 18 and 65 shows that the pandemic’s effects on mortality have been greatest among essential workers, particularly those in the food/agriculture, transportation/logistics, facilities, and manufacturing sectors. Such workers experienced an increased risk of mortality of greater than 20% during the pandemic, with an increased risk of greater than 40% during the first two full months of the state’s reopening. Excess mortality in high-risk occupational sectors was evident in analyses stratified by race/ethnicity, especially for Latino, Black, and Asian workers.

Our findings are consistent with a small but growing body of literature demonstrating occupational risks for SARS-CoV-2 infection. For example, a study of the UK Biobank cohort found that essential workers, particularly healthcare workers, had high risks for COVID-19 [10]. Similarly, numerous studies have documented SARS-CoV-2 infection among healthcare workers [11]. Our study, however, is unique in examining excess mortality and multiple occupational sectors. Though our work is in agreement with prior studies in finding pandemic-related risks among healthcare workers [11], it suggests that the risks are even higher in other sectors, such as food/agriculture and transportation/logistics.

This study is also among the first to examine deaths by both occupation and race/ethnicity. Occupational exposures have been postulated as an important contributor for disparities in excess mortality by race ethnicity, particularly because certain occupations require in-person work [4]. Though we tended to find the largest relative increases in mortality in each racial/ethnic group in the food/agriculture and transportation/logistics sectors, there was variation across race/ethnicity. For example, among Asians, the largest RR was in the health/emergency sector, even though the relative risk increases in that sector were relatively low among other racial/ethnic groups. Such differences may reflect cross-sector differences in demographics. There are, for example, a large number of Latinos who work in meat-processing facilities [12], consistent with data that show that Latinos make up a large proportion of COVID-19 cases in such settings [13]. Similarly, the large RR among Asians in the health/emergency sector could be due to the relatively large number of Filipino Americans in nursing professions [14]. During the pandemic in particular, such disproportionate representation may easily lead to cross-race variability in risk. A recent study found, for example, that Black workers are more likely to be employed in occupations that frequently require close proximity to others [15]. Inequalities in risk may be exacerbated by underlying structural inequities, such as immigration status or poverty [16].

Though non-occupational risk factors may be relevant, it is clear that eliminating COVID-19 will require addressing occupational risks. In-person essential workers are unique in that they are not protected by shelter-in-place policies. Indeed, our study shows that excess mortality rose sharply in the food/agriculture sector during the state’s first shelter-in-place period, from late March through May; these increases were not seen among those working in non-essential sectors. Complementary policies are necessary to protect those who cannot work from home. These can and should include: free personal protective equipment, clearly defined and strongly enforced safety protocols, easily accessible testing, generous sick policies, and appropriate responses to workplace safety violations. As jurisdictions struggle with difficult decisions regarding vaccine distribution, our findings offer a clear point of clarity: vaccination programs prioritizing workers in sectors such as food/agriculture are likely to have disproportionately large benefits for reducing COVID-19 mortality.

We acknowledge limitations to the study, including misclassification of occupation in death certificates due to coarse categories or inaccurate reports. The decedent’s primary occupation is typically reported by the next of kin who may not be able to precisely describe the work. The primary occupation, which is reported on the death certificate, may not match the most recent occupation, which is more likely to drive occupational risk. These limitations would in general attenuate apparent differences across occupational sectors but are unlikely to account for our primary results.

Our study places a powerful lens on the unjust impact of the COVID-19 pandemic on mortality of working age adults in different occupations. Our analysis is among the first to identify non-healthcare in-person essential work, such as food and agriculture, as a predictor of pandemic-related mortality. Essential workers—especially those in the food/agriculture, transportation/logistics, facilities, and manufacturing sectors—face increased risks for pandemic-related mortality. Shutdown policies by definition do not protect essential workers and must be complemented with workplace modifications and prioritized vaccine distribution. If indeed these workers are essential, we must be swift and decisive in enacting measures that will treat their lives as such.

## Data Availability

Data are not publicly available.

